# Cold plasma for SARS-CoV-2 Inactivation

**DOI:** 10.1101/2020.09.17.20192872

**Authors:** Zhitong Chen, Gustavo Garcia, Vaithilingaraja Arumugaswami, Richard E. Wirz

## Abstract

SARS-CoV-2 infectious virions are viable on various surfaces (e.g., plastic, metals, cardboard) for several hours. This presents a transmission cycle for human infection that can be broken by developing new inactivation approaches. We employed an efficient cold atmospheric plasma (CAP) with argon feed gas to inactivate SARS-CoV-2 on various surfaces including plastic, metal, cardboard, basketball composite leather, football leather, and baseball leather. These results demonstrate the great potential of CAP as a safe and effective means to prevent virus transmission and infections.

---

Severe acute respiratory syndrome coronavirus 2 (SARS-CoV-2) has caused a once-in-a-century pandemic and studies have shown that the infectious virions are viable on various surfaces (e.g., plastic, metals, cardboard) for several hours.^1–4^ Surface contamination presents a great risk of transmitting SARS-CoV-2 between people and it is critical to break the transmission cycle by developing new inactivation approaches.

Plasma is one of the four fundamental states of matter (i.e., solid, liquid, gas, and plasma) and was so named since the charged species that comprise the plasma can behave somewhat similarly to the cellular components of blood that are bound by blood plasma. Cold atmospheric plasma (CAP) operating at atmospheric pressure and room temperature is useful for safely treating contaminated surfaces and can treat smooth and highly-featured surfaces. The efficacy of CAP is due to its many components, such as reactive oxygen and nitrogen species (RONS) (Fig. 1 A), which exhibit favorable behavior for biomedical applications.^5–7^ The biophysical details are discussed in our previous paper.^8^

**Figure 1.**
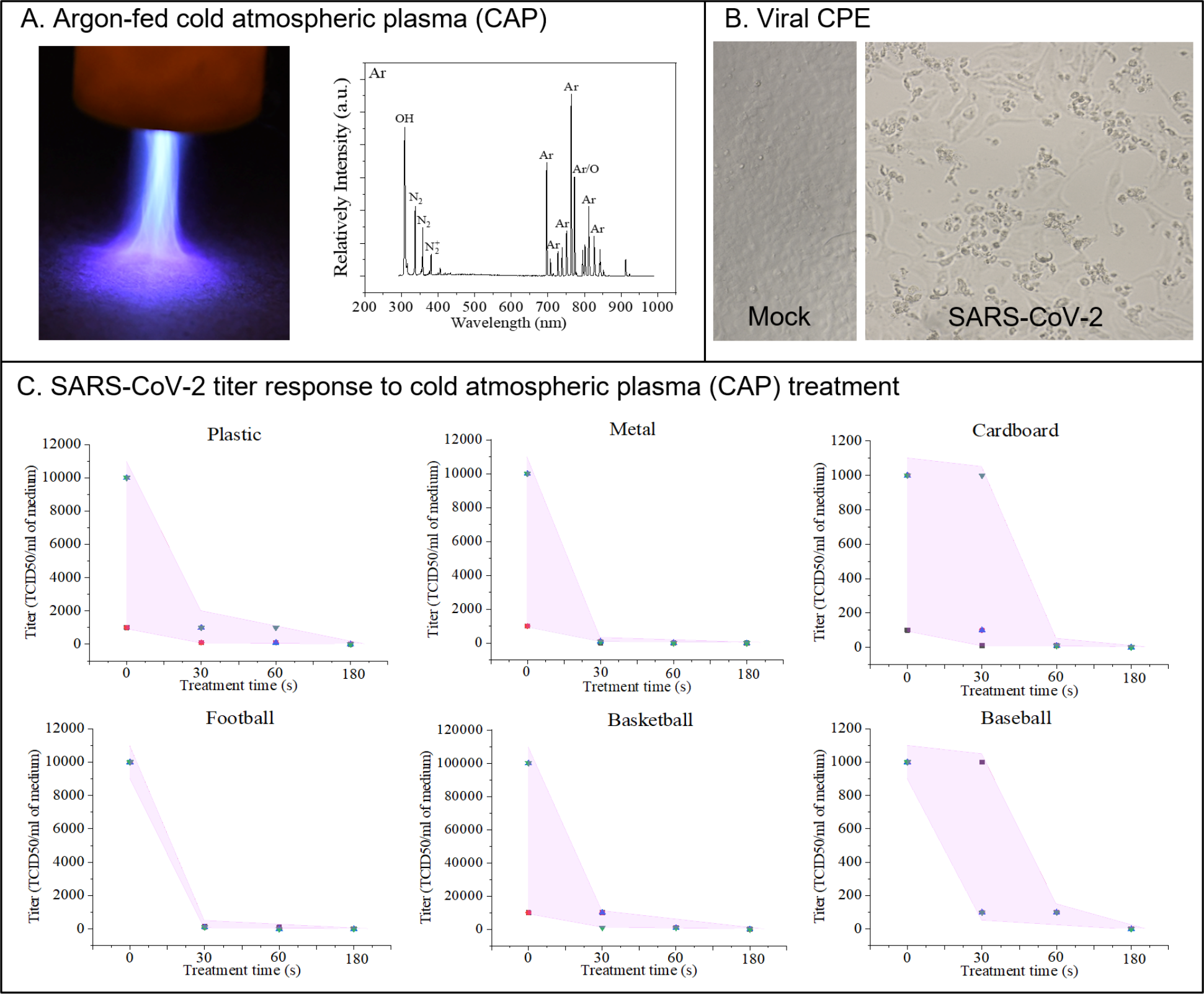
Ar-fed CAP disinfecting SARS-CoV-2. Panel A shows Ar-fed CAP treatment of a plastic surface and the optical emission spectrum of reactive oxygen and nitrogen species (RONS) (Exposure: 250 ms). Panel B shows a Bright-field image of SARS-CoV-2 infected Vero-E6 cells showing viral cytopathic effect (CPE). Uninfected (Mock) cells included as control. Panel C shows SARS-CoV-2 titer response to CAP treatment times of 0, 30, 60, and 180 seconds on surfaces of plastic, metal, cardboard, leather football, composite leather basketball, and leather baseball. Error bar in each graph is shown as shaded area.

We employed argon(Ar)-fed CAP treatments to inactivate SARS-CoV-2 on various surfaces including plastic, metal, cardboard, basketball composite leather, football leather, and baseball leather. The CAP device consists of a two-electrode assembly with a powered needle electrode and a grounded outer ring electrode, which were connected to a high voltage transformer. The custom body was developed with a 3D printer (LulzBot TAZ 6) at UCLA.^9^ Discharge voltage for both feeding gases was 16.6 kV (peak-peak) at 12.5 kHz frequency. The RMS input power for the device is approximately 12 W. SARS-CoV-2 infected Vero-E6 cells show a viral cytopathic effect (Fig. 1B). First and foremost, this is the first-ever demonstration of plasma inactivation of SARSCoV-2 and is a significant milestone for the biotech community.

We observed that Ar-fed CAP treatment inactivated all SARS-CoV-2 on the six surfaces in less than 180 seconds (Fig. 1C) and specifically metal and the leather football surfaces exhibited decontamination by 30 seconds of exposure. Most data points on the plastic surface showed virus inactivation for 30 and 60 second treatments. Cardboard and basketball surfaces exhibited effective virus inactivation for a 60 second treatment, while few data points exhibit these effects for 30 second treatments. Additional testing showed similar virus inactivation for cotton cloth material from face masks.

Initial testing was performed with helium (He)-fed CAP. Both He-fed and Ar-fed plasmas operated at atmospheric pressure and room temperature (Fig. 2). Unlike Ar-fed plasma, He-fed plasma did not disinfect all SARS-CoV-2 on metal and plastic surfaces even at 300 seconds (Fig. 3). This is likely due to the He-fed plasmas much lower RONS concentrations when compared to Ar-fed plasma for the same operating conditions (compare Fig. 1A for Ar-fed to Fig. 4 for He-fed).

**Figure 2.**
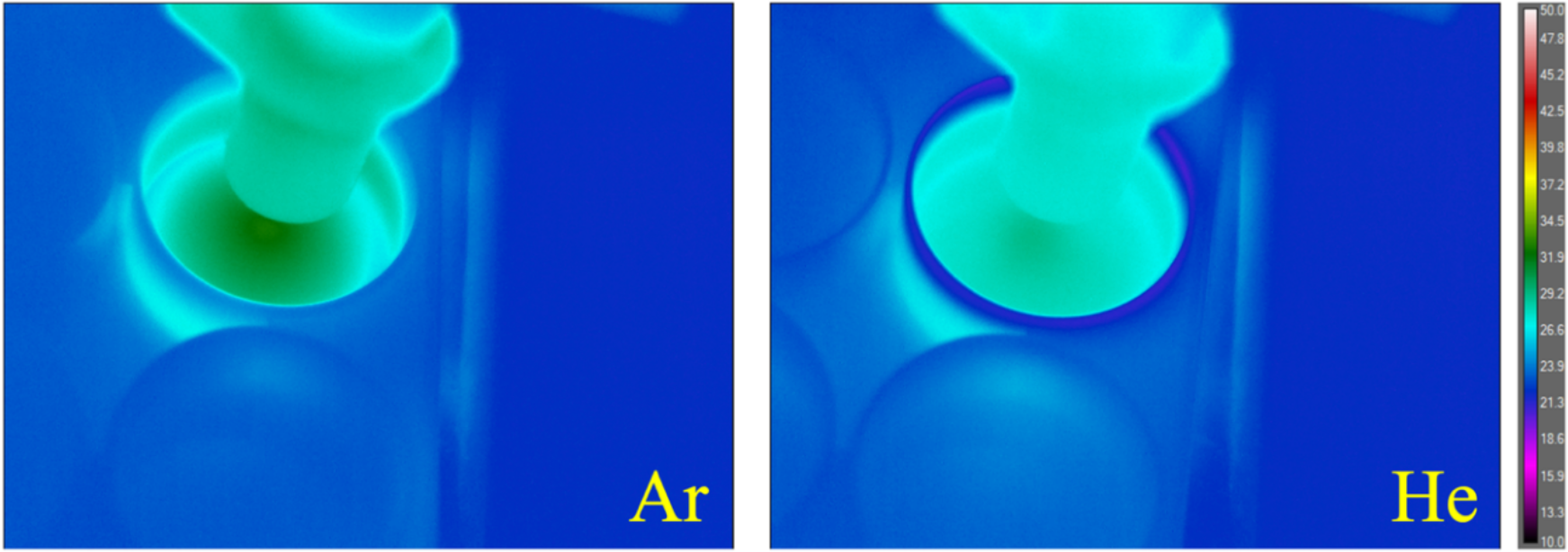
Temperature distribution of Ar and He plasma-treated 6-well plate. The highest temperature for each subject was found at the center of the circular thermal gradient immediately beneath the plasma discharge. The center temperature for the Ar plasma treated surface was approximately 32 °C and for the He plasma treatment was approximately 29 °C.

**Figure 3.**
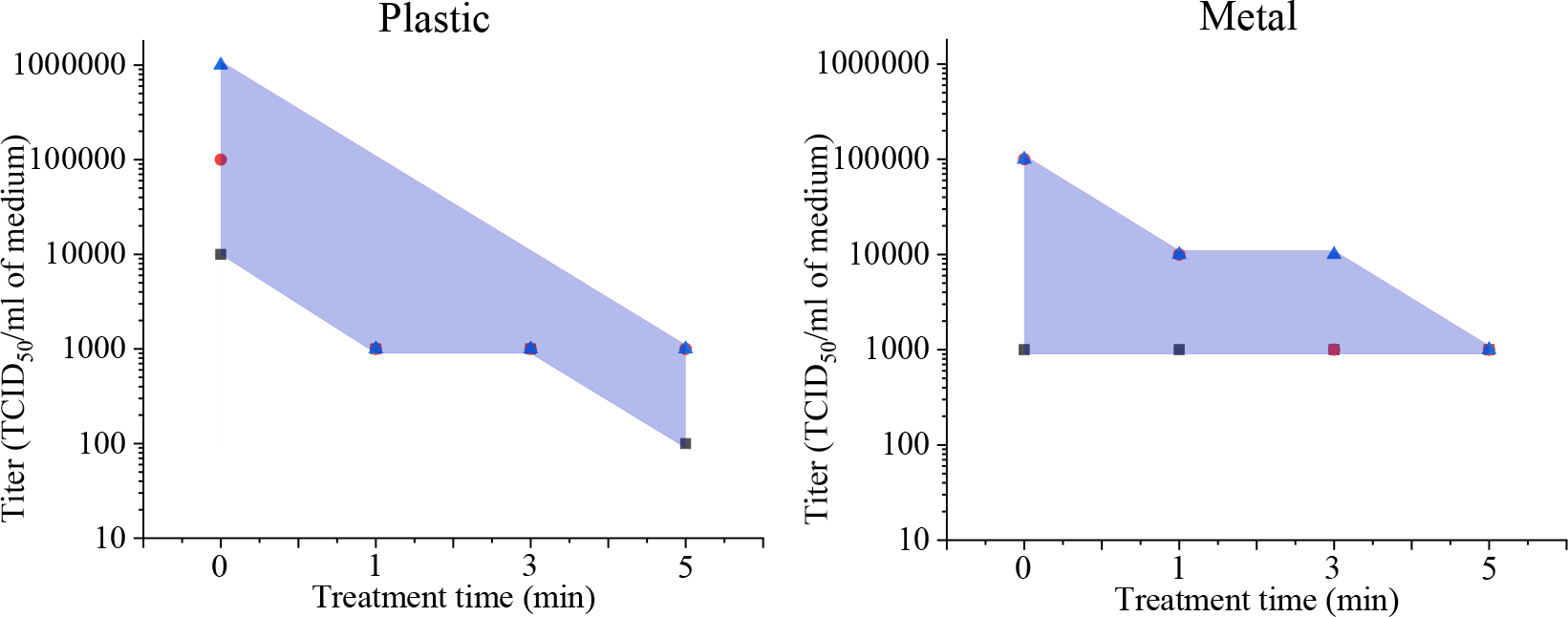
He-fed CAP disinfecting SARS-CoV-2. He-fed CAP disinfecting SARS-CoV-2 on plastic and metal surfaces. Error bar in each graph is shown as shaded area.

**Figure 4.**
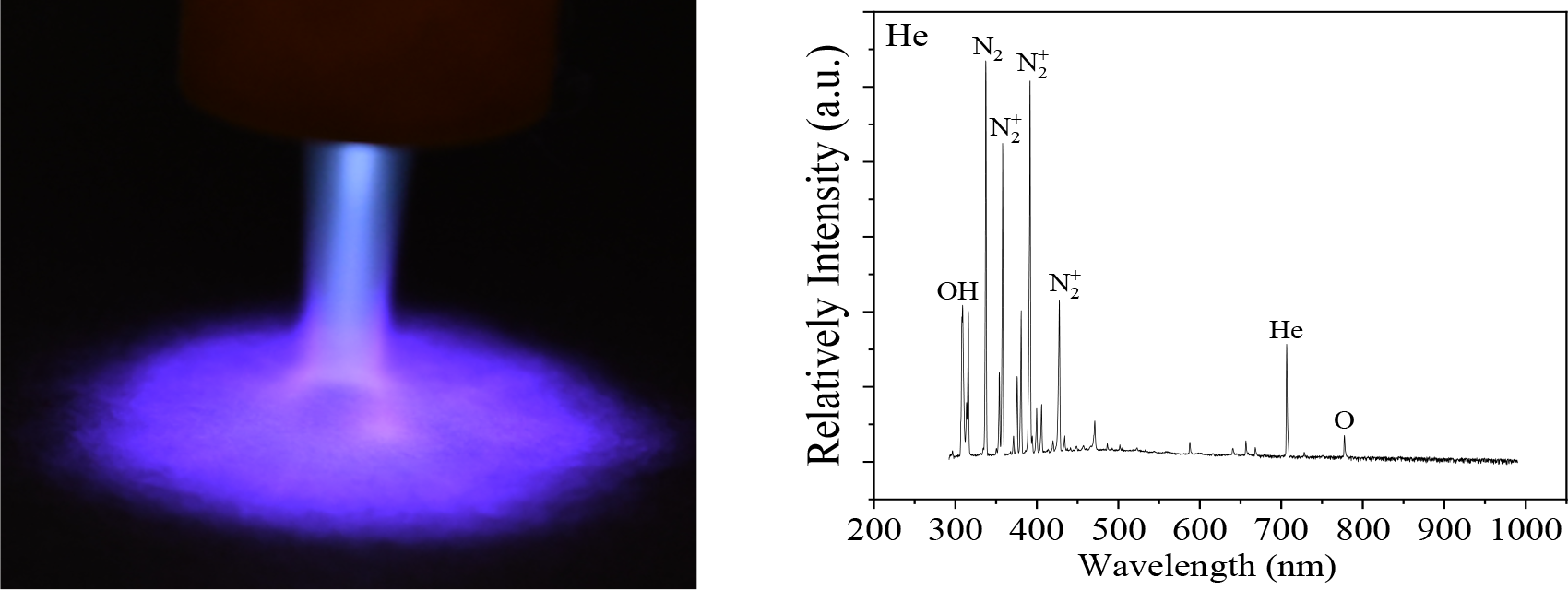
He-fed CAP and its spectrum. Left: He-fed plasma jet treating surface with plume spreading across the surface. Right: The He-fed plasma jet was detected by the optical emission spectrum and contained reactive oxygen and nitrogen species (RONS) (Exposure: 5000 ms).

## Data Availability

Please contact with authors for data.

## Overall

Ar-fed CAP treatment was shown to effectively inactivate SARS-CoV-2 on a wide range of surfaces that people touch and therefore has great potential as a safe and effective means to prevent virus transmission and infections. Since cold plasma is significantly safer than most other treatment methods such as alcohol, UV radiation, and the like; this work opens a wide range of opportunities for the scientific, engineering, and medical communities.

## Acknowledgements

This research was partially funded by the Air Force Office of Scientific Research (FA9550-14-10317) UCLA Subaward (No. 60796566-114411) to R.E.W and UCLA DGSOM and Broad Stem Cell Research Center institutional award (OCRC #20–1) to V.A.

## Methods and Materials

SARS-Related Coronavirus 2 (SARS-CoV-2), Isolate USA-WA1/2020, was received from BEI Resources of National Institute of Allergy and Infectious Diseases (NIAID). SARS-CoV-2 live culture studies were conducted in UCLA BSL3 high-containment facility. SARS-CoV-2 was passaged twice in Vero-E6 cells and viral stocks were prepared in 1 ml aliquots and stored at - 80oC. Vero-E6 cells were used for measuring virus titer by TCID50 assay. Cells were cultured in DMEM growth media containing 10% fetal bovine serum (FBS), 2 mM L-glutamine, penicillin (100 units/ml), streptomycin (100 units/ml), and 10 mM HEPES. Cells were incubated at 37 °C with 5% CO_2_.

Various surfaces were treated with SARS-CoV-2 at 2 × 10^5^ PFU in a 25 ul volume. Virus contaminated surfaces were exposed to Helium or Argon plasma in various durations. Surfaces treated with virus, but not exposed to plasma were included as control. Infectious SARS-CoV-2 were recovered from surfaces by adding 100 ul of DMEM growth media. The titer of recovered SARS-CoV-2 was assessed in Vero-E6 cells in a 96-well format. Each recovered sample was subjected to 10-fold serial dilution and 100 ul of diluted viral inoculum was added to each well. At 3 to 4 days post infection, wells were examined for presence viral cytopathic effect (CPE). At the lowest viral dilution, wells negative for CPE were identified and included for calculating virus titer for each sample.

Optical emission spectroscopy for the plasma was performed with a fiber-coupled optical spectrometer (LR1-ASEQ Instruments), with a range of wavelength 300 –1000 nm. These measurements detected the CAP-generated ROS and RNS (such as nitric oxide [NO], nitrogen cation [N_2^+^_], atomic oxygen [O], and hydroxyl radicals [•OH]). The optical probe was placed at a radial distance of 10 mm from the center of the plasma jet. Data were collected with an exposure time of 250 ms and 5000 ms for Ar and He, respectively.

Thermal measurements of the subject were made with a tripod-mounted long-wavelength infrared camera (FLIR A655sc) from a distance of approximately 15 cm diagonally above the subject; the relative position of the camera to the subject remained consistent for all tests. Frame sequences for a 60-second exposure were recorded using Research IR 4.40 with individual frames extracted as needed after recording. A linear scale manually configured from 10 °C to 60 °C was selected for all images to allow for a sufficient dynamic range for both Ar and He tests.

